# A Cross-sectional Comparison of Utilization of Medical Crowdfunding for Global Health

**DOI:** 10.1101/2021.10.05.21264582

**Authors:** Andie Siemens, Samantha Rustja, R. Christopher Doiron, D. Robert Siemens

## Abstract

The online solicitation of public donations has become an important financing option for health care expenses, intensified by increasing costs and deficits of universal public systems (1). With growing internet access and success of the largest social crowdfunding platform, GoFundMe, online appeals for medical causes have grown significantly over the last decade in low-to high-income countries. The purpose of this study was to qualitatively describe the use of GoFundMe as a crowdfunding platform for global health initiatives given its supremacy in the social crowdfunding market. Three different cohorts (n=100 each) of online solicitation were examined as a cross-section comparing global health appeals to those for personal health care and animal activism. Variables included the purpose for crowdfunding, the characteristics of beneficiaries and campaigns, and the factors associated with funding success. Our cross-sectional review found that global health campaigns were focused on voluntourism opportunities compared to more specific, individualized appeals for those in need. Global health campaigns appeared to be the least ambitious and generally the least successful of those reviewed. Grouping the most and least successful campaigns between the different cohorts, global health appears to be more successful when targeting a larger population to donate smaller amounts of money and relying on sharing via social media. We suggest that compared to online solicitation for personal health and animal activism objectives, crowdfunding on GoFundMe has unrealized potential as a tool for global health initiatives. More work should be conducted using different crowdfunding platforms and a more longitudinal review in order to expand on these findings and their implications on health care provision in the countries examined. Furthermore, future inquiry is needed to understand the social and ethical implications of online solicitation for global health endeavors in order to inform policy and promote discussion around equity and accessibility.

## Introduction

There is a significant discrepancy between countries in healthcare infrastructure. Low- and middle-income countries (LMICs) tend to have more difficulty providing medical care to its citizens for a wide range of reasons, including restricted access to modern medications due to high costs and uneven distribution of medical professionals (2). As one example, Africa has a critical healthcare shortage with a ratio of 2.3 doctors per 1000 citizens compared to the Americas’ ratio of 24.8(2). Online solicitation of public donations has become an important financing option for health care expenses, intensified by increasing costs in jurisdictions without universal public systems including the United States (US) (3). These online appeals for personal medical causes have grown significantly over the last decade in developed countries although the utility of medical crowdfunding in different countries vary (4). In high-income countries (HICs) with universal healthcare such as Canada, there is less concern about personal expenses to cover costs of medical treatments opening up an opportunity for global altruism and resource sharing. Crowdfunding websites such as GoFundMe, the largest social crowdfunding platform in the world,(5,6) as a global health tool could theoretically act as a bridge between HICs and LMICs by connecting individuals capable of donating with individuals in need of funding through highly personal charity opportunities.

Such online crowdfunding opportunities for global health activity could act as an alternative or supplement to other types of foreign aid, including voluntourism (7), as well as governmental or non-governmental organization (NGO) activities. Developed countries with universal health care systems such as Canada have long histories of providing various types of global health relief to countries in need and has largely contributed to progression in global health (8). Canada is 3^rd^ out of the Group of Seven in terms of development assistance with respect to health, donating $1.25 billion CAD in 2016 (8). Governmental and large organization global health initiatives benefit from their scale and ability to create coalitions: contributing to research questions, priority setting, and fostering relevance and effectiveness of initiatives through knowledge translation. This being said, these initiatives are often broader in scope with individual donors less likely to know the exact use of funds. In contrast, online crowdfunding sites allow individuals to pick and choose causes that are meaningful to them to solve specific health issues. It also allows the public to follow the change their donation has made and marketing through social media to increases engagement.

This current study’s objective is to qualitatively describe the state of global health initiatives on the crowdfunding site GoFundMe, examining the campaigns’ composition, goals, and activity. In order to contextualize the observations we report on the relative success of global health campaigns compared to those fundraising for personal medical situations (referred to as personal health) and for non-human, domestic and wild animal welfare (referred to as animal activism). We selected GoFundMe as an ideal environment to study given its market supremacy and specifically accessed campaigns from the platform in Canada with its relatively large market but also its potentially unique use of such online solicitation platforms for medical care given its universal health care system (4).

## Methods

We sought to explore crowdfunding campaigns for global health endeavors hosted by the website GoFundMe (https://www.gofundme.com/) and compare these to similar drives on the website: personal health and animal activism campaigns. An *a priori* decision was to review 100 campaigns for each topic and assess whether saturation of pre-determined descriptive variables as well as any novel concepts regarding the campaigns had occurred. For the personal health cohort, the first 50 campaigns from the website’s “medical” category were first captured for analysis. Additionally, the first 25 campaigns corresponding respectively to the general search terms “help” and “support” were used to capture campaigns external to GoFundMe’s “medical” category. Animal activism campaigns were captured in a similar fashion – the first 50 campaigns from the website’s “animal” category followed by the first 25 campaigns from respective general search terms “wildlife” and “conservation.” GoFundMe provides no explicit “global health” category in their search function so the first 100 campaigns from respective general search terms “global health” and “medical mission trip” were used to capture global health campaigns for analysis. All searches were performed on GoFundMe’s publicly accessible website from Canada between September, 2019–September, 2020.

Descriptive variables captured during the campaign assessments included the type of recipient as an individual or organization; for the personal health and global health cohorts the recipient’s demographics were further categorized as pediatric versus adult while the animal activism cohort was described in terms of type of recipient animal. The campaign organizer was identified as the patient, friend/family member, or an organization. Geographic origin of the campaign organizer and beneficiary was captured by country. The mission of care for the campaign and specific use of the funds was also summarized. Furthermore, several campaign descriptors were denoted including campaign images, named religious/spiritual affiliations, environmental impact, and the presence of low resource group(s). Descriptive variables collected were then sorted by a second author (AS, SR) with any disagreements in sorting adjudicated by a third party (DRS).

Quantitative variables were captured, including campaign goal amount (CAD$), amount raised (CAD$), number of donors, average donation amount, and the number of shares (a function provided by GoFundMe to directly share to Facebook, Twitter, WhatsApp, email, text, or copy URL). Medians and interquartile ranges were calculated for all quantitative variables. A campaign success index (CSI) was calculated as the percent of the campaign goal raised divided by number of days from when the campaign was posted to the final day of data collection (September 15, 2020) and multiplied by 1000. Campaigns with goal values of $1 or no goal and outlier campaigns that raised >1000% of their fund-raising goal were excluded from CSI calculations. A descriptive subset analysis comparing the top 20 campaigns to the bottom 20 campaigns based on CSI rankings was performed to examine factors relating to campaign success.

## Results

### Campaign recipient and organizer demographics

The majority (83/100; 83%) of recipients of the global health crowdfunding campaigns were directed to specific individuals, similar to those campaigns for personal health issues (98%) (Table 1). Comparatively, 65% of animal activism campaigns directed their funding to named organizations. As expected, the demographics for the identified recipient of personal health campaigns were well described (70% were for adult patients) whereas the majority (85%) of campaigns for global health had little demographic information for the recipient included within the campaign. Recipients of the of animal activism campaigns were equally divided between domestic and wild animals (44% and 50%, respectively).

**Table 1.**
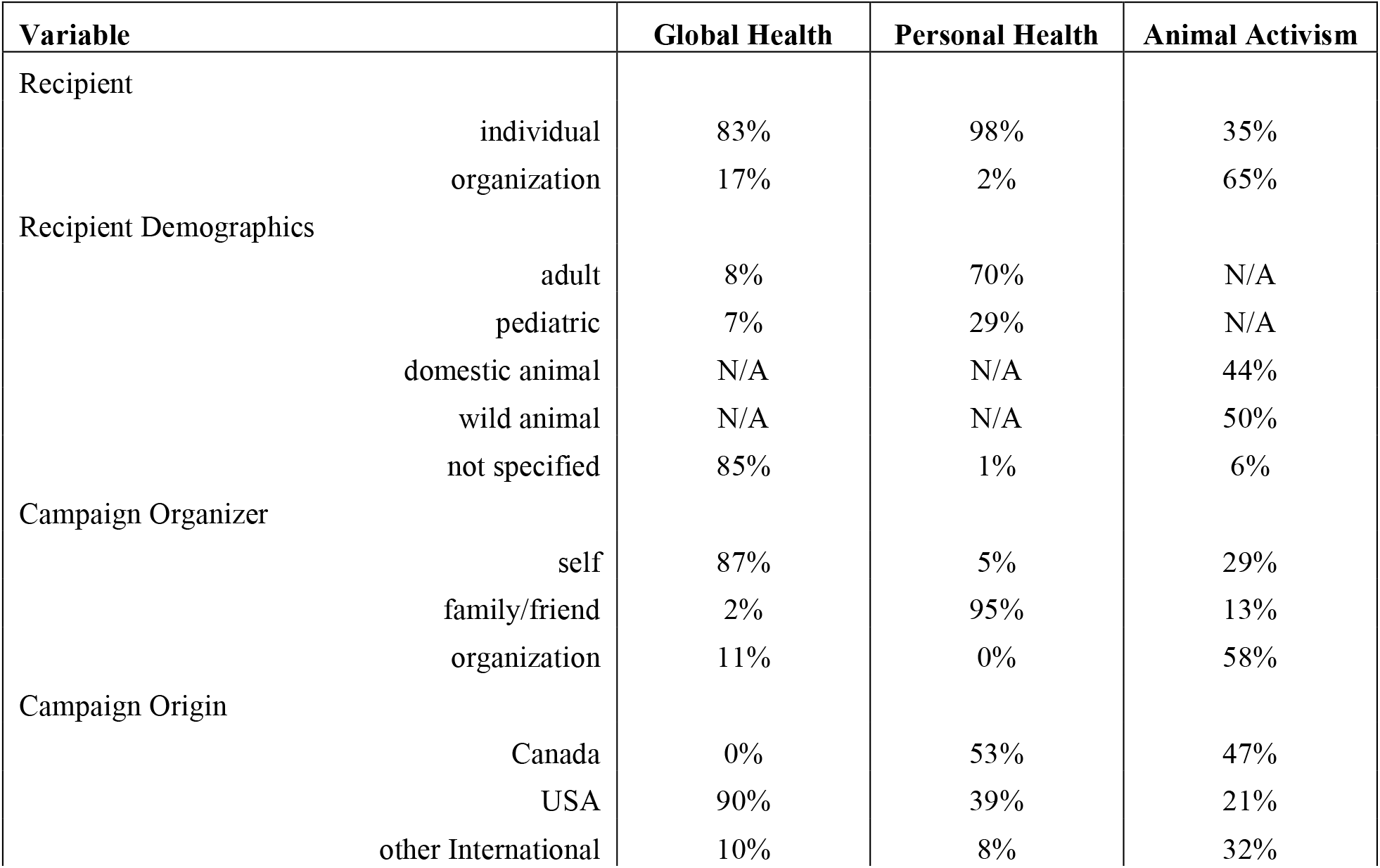
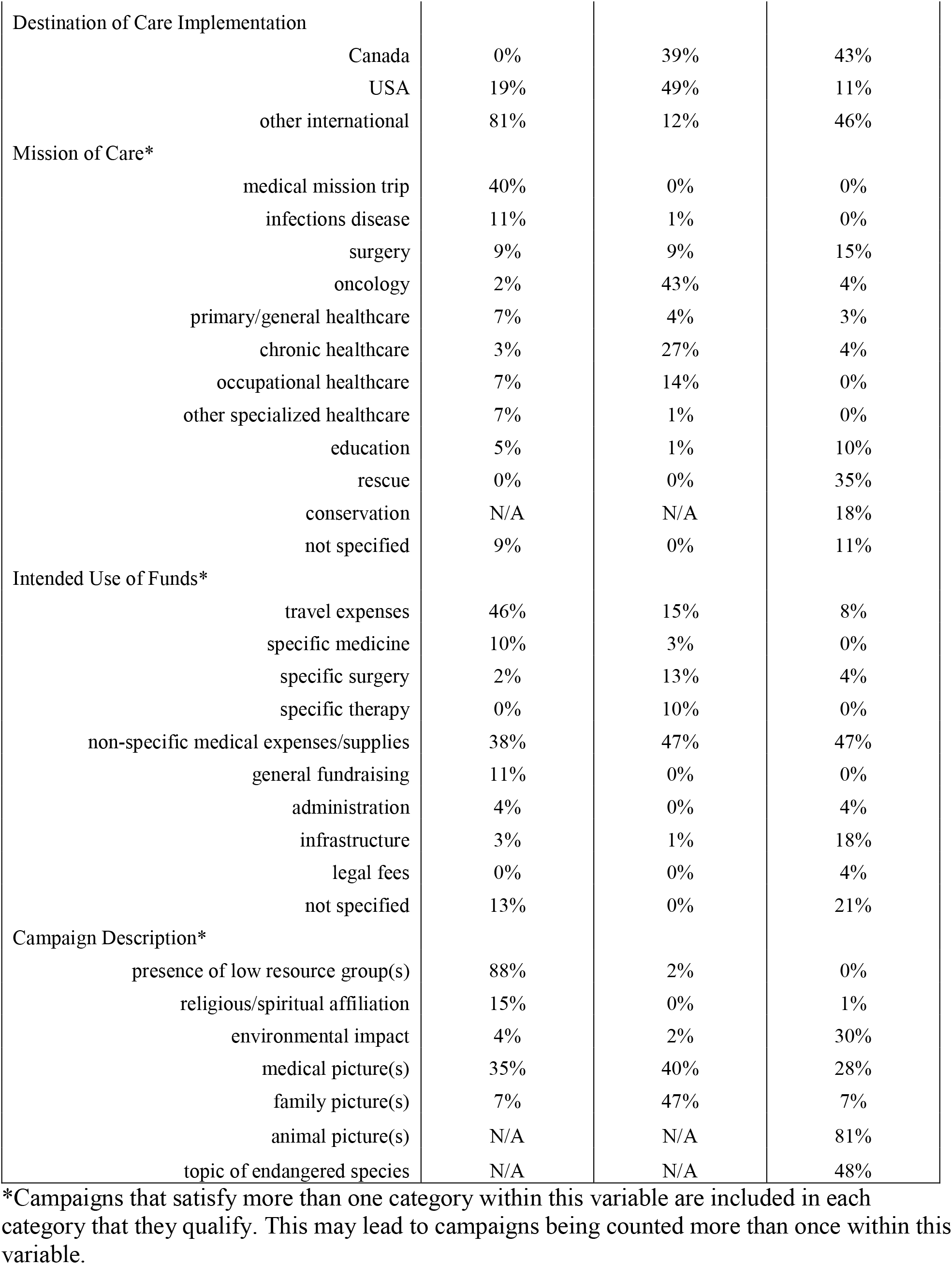
Global health, personal health, and animal activism campaign composition.

| Variable | Global Health | Personal Health | Animal Activism |
| --- | --- | --- | --- |
| Recipient |  |  |  |
| individual | 83% | 98% | 35% |
| organization | 17% | 2% | 65% |
| Recipient Demographics |  |  |  |
| adult | 8% | 70% | N/A |
| pediatric | 7% | 29% | N/A |
| domestic animal | N/A | N/A | 44% |
| wild animal | N/A | N/A | 50% |
| not specified | 85% | 1% | 6% |
| Campaign Organizer |  |  |  |
| self | 87% | 5% | 29% |
| family/friend | 2% | 95% | 13% |
| organization | 11% | 0% | 58% |
| Campaign Origin |  |  |  |
| Canada | 0% | 53% | 47% |
| USA | 90% | 39% | 21% |
| other International | 10% | 8% | 32% |

|  |  |  |  |  |
| --- | --- | --- | --- | --- |
| Destination of Care Implementation |  |  |  |  |
|  | Canada | 0% | 39% | 43% |
|  | USA | 19% | 49% | 11% |
|  | other international | 81% | 12% | 46% |
| Mission of Care* |  |  |  |  |
|  | medical mission trip | 40% | 0% | 0% |
|  | infections disease | 11% | 1% | 0% |
|  | surgery | 9% | 9% | 15% |
|  | oncology | 2% | 43% | 4% |
|  | primary/general healthcare | 7% | 4% | 3% |
|  | chronic healthcare | 3% | 27% | 4% |
|  | occupational healthcare | 7% | 14% | 0% |
|  | other specialized healthcare | 7% | 1% | 0% |
|  | education | 5% | 1% | 10% |
|  | rescue | 0% | 0% | 35% |
|  | conservation | N/A | N/A | 18% |
|  | not specified | 9% | 0% | 11% |
| Intended Use of Funds* |  |  |  |  |
|  | travel expenses | 46% | 15% | 8% |
|  | specific medicine | 10% | 3% | 0% |
|  | specific surgery | 2% | 13% | 4% |
|  | specific therapy | 0% | 10% | 0% |
|  | non-specific medical expenses/supplies | 38% | 47% | 47% |
|  | general fundraising | 11% | 0% | 0% |
|  | administration | 4% | 0% | 4% |
|  | infrastructure | 3% | 1% | 18% |
|  | legal fees | 0% | 0% | 4% |
|  | not specified | 13% | 0% | 21% |
| Campaign Description* |  |  |  |  |
|  | presence of low resource group(s) | 88% | 2% | 0% |
|  | religious/spiritual affiliation | 15% | 0% | 1% |
|  | environmental impact | 4% | 2% | 30% |
|  | medical picture(s) | 35% | 40% | 28% |
|  | family picture(s) | 7% | 47% | 7% |
|  | animal picture(s) | N/A | N/A | 81% |
|  | topic of endangered species | N/A | N/A | 48% |

\*Campaigns that satisfy more than one category within this variable are included in each category that they qualify. This may lead to campaigns being counted more than once within this variable.

The global health campaigns were most commonly self-organized by the funding recipient (87%) while the majority of personal health campaigns were organized by a friend or family member (95%). Of the animal activism campaigns, 58% originated from the organizations that were also the beneficiary of the crowdfunding.

Interestingly the majority (90%) of global health campaigns captured originated from the US with none originating from Canada. As expected by accessing the platform from a Canadian location, the majority of the personal health campaigns identified originated from Canada (53%) or US (39%). The animal activism campaign origins spread between Canada (47%), US (21%), and other countries (32%). None of the identified global health campaigns stated plans for implementation within Canada with the majority (81%) of campaigns listing international destinations for their programs of care and the remainder destined for the US (19%). The majority of personal health campaigns planned to implement care in Canada (39%) or the US (49%) while the animal activism campaigns planned to implement action in Canada (43%) or an international destination (46%).

### Mission of Care and Intended Use of Funds

A large proportion (40%) of the global health campaigns were dedicated to unspecified mission trips while most personal health campaigns had specific descriptions of the care needed including oncology (43%), chronic care (27%), and occupational health (14%). Animal activism campaigns were often focused for rescue purposes (35%). Most of the global health drives stated their intended use of funds as being for travel expenses (46%) or for non-specific medical expenses/supplies (38%). Global health campaigns not requiring travel expenses dedicated a large proportion of their funding to non-specific medical expenses/supplies, as was similarly apparent in personal health and animal activism campaigns (Table 1).

There was a religious or spirituality affiliation in 15% of the global health cohort, while none of the identified personal health campaigns and only one of the animal activism campaigns provided religious motivation or inspiration. Natural disasters, such as wildfires, were commonly stated as motivators for animal activism campaigns (30%). The majority (88%) of global health campaigns implied a low resource group in need as a motivator for the campaign. Personal health campaigns included images depicting medical topics or family members in 87% of the time, compared to 42% in global health campaigns and 35% of the animal activism cohort.

### Campaign Goals and Activity Details

Global health campaigns had the lowest fundraising goals with a median of $3,000 (Interquartile range (IQR): $2,000-$8,500). Personal health campaigns had the largest fundraising goals followed by animal activism with a median of $52,500 (IQR: $20,000-$150,000) and $15,000 (IQR: $6,516-$50,000), respectively with a substantial degree of variation among them (Fig 1A). The amount of funds raised within the global health campaigns were also the lowest with a median of $2,150 (IQR: $1,600-$4,480), compared to $32,500 (IQR:$14,250-$74,750 and $8,208 (IQR:$6,294-$19,956) for personal health and animal activism respectively (Fig 1B). In a similar trend, the number of donors [median 30 (IQR: 19-64)] to the global health campaigns were considerably lower than the other campaigns (Fig 1C). The average per donation amount was similar for global health compared to the other campaigns however, the personal health and animal activism campaigns had many more donors leading to the higher total dollar amounts raised (Fig 1D). Similarly, the number of campaign shares to social media and content sharing platforms (email, text, and copy URL) for global health campaigns was the lowest with a median of 155 (IQR: 88-333) compared to the personal health [1,000 (IQR: 527-2700)] and animal activism campaigns [391 (IQR: 136-1,100)] (Fig 1E).

**Fig 1.**
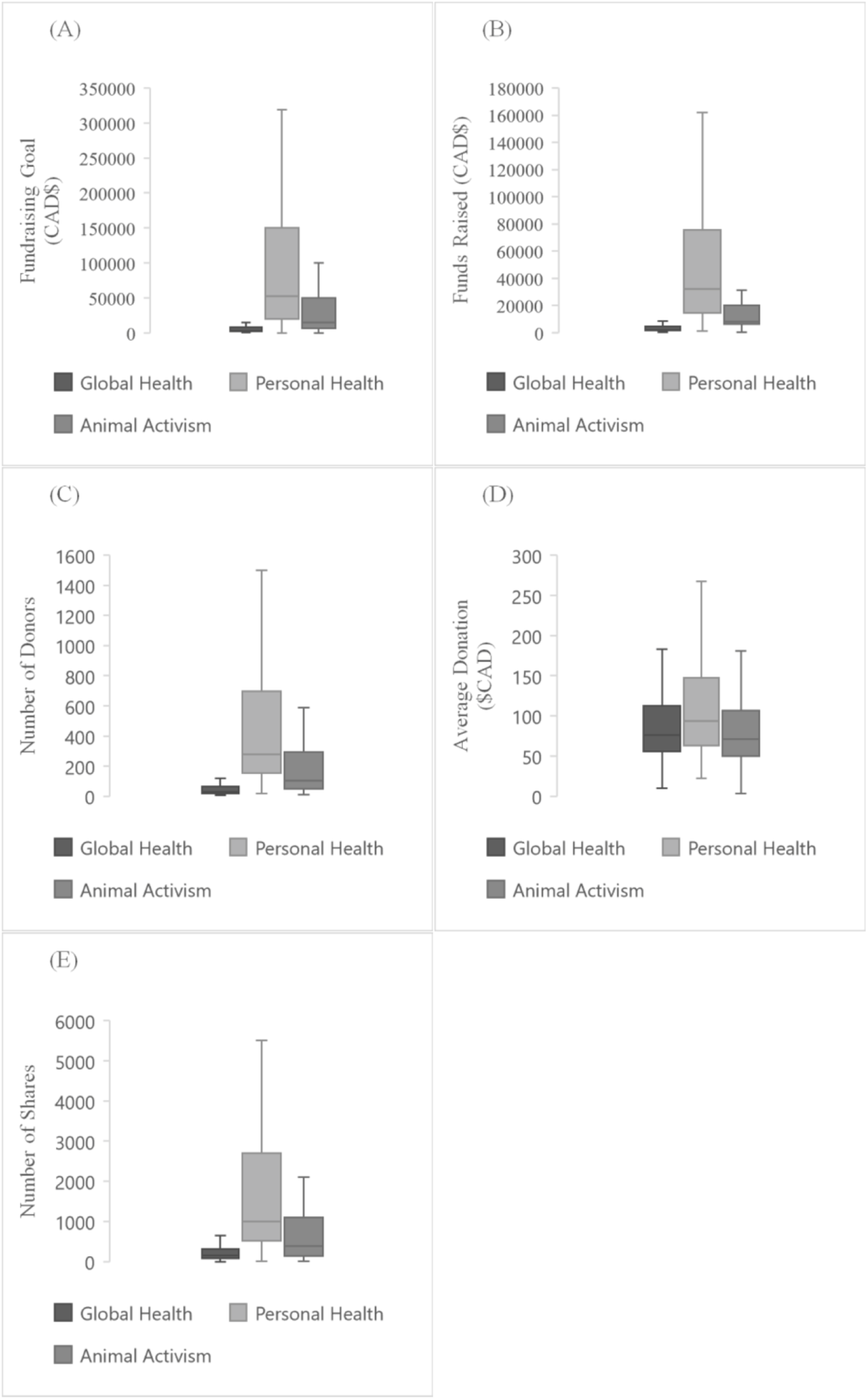
Comparison of campaign composition in global health, personal health, and animal activism. n=100 for each cohort. (A) Fundraising goal in CAD$. (B) Amount of funds raised in CAD$. (E) Number of donors donating to the campaigns. (D) Average donation amount in CAD$ calculated from the funds raised divided by the number of donors campaign for global health, personal health, and animal activism campaigns. (E) Number of shares of the campaigns through the GoFundMe share function to social media and content sharing platforms.

### Campaign Success

Based on the calculated campaign success index, personal health campaigns were the most successful with a median CSI of 2.51 (IQR: 1.39-3.95), followed by animal activism campaigns 2.05 (IQR: 0.80-3.17). The global health campaigns were least successful at 1.24 (IQR: 0.69-2.73) (Fig 2).

**Fig 2.**
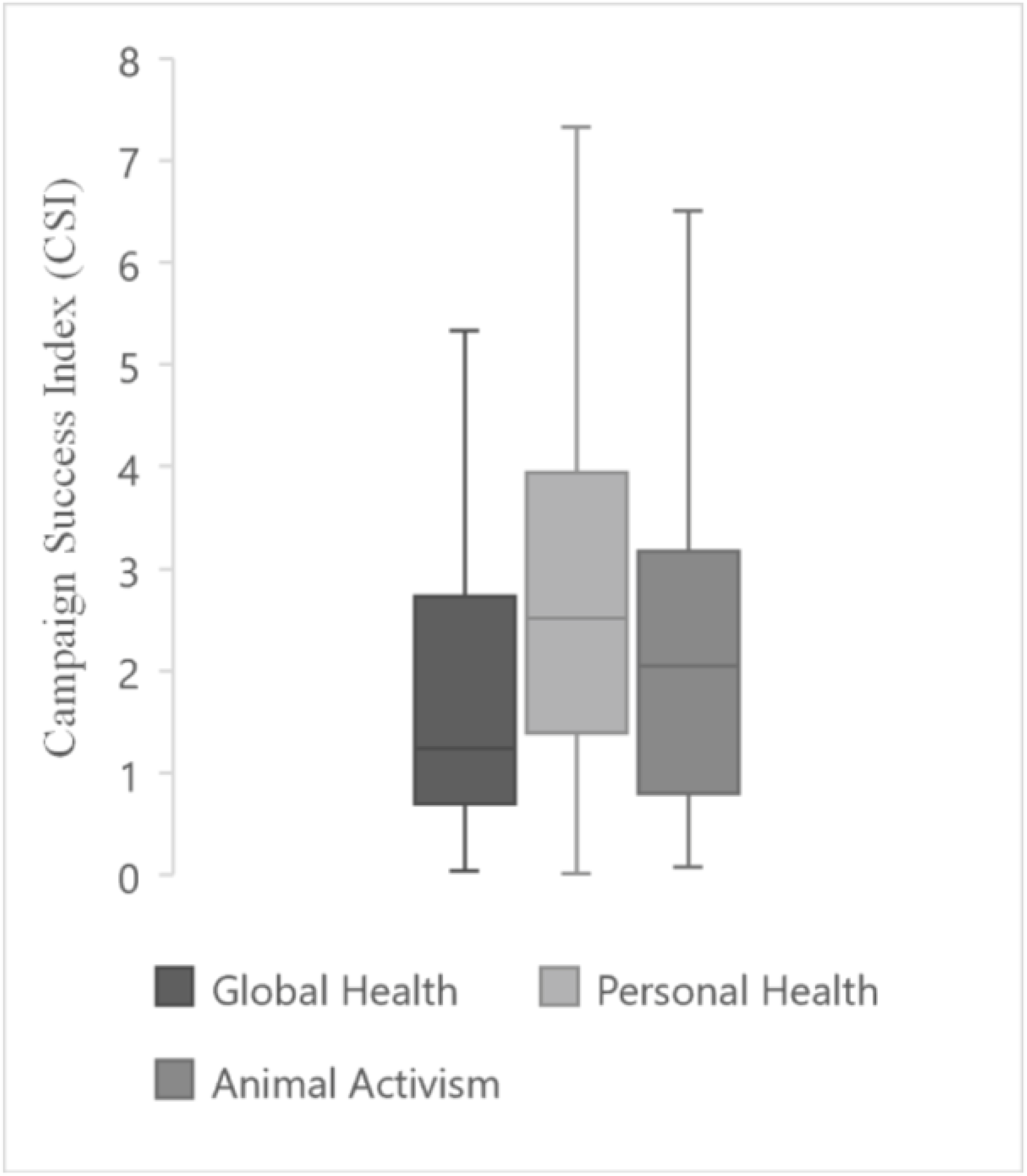
Campaign success index (CSI) for global health, personal health, and animal activism campaigns. CSI calculated as the percent of the campaign goal raised divided by number of days from when the campaigned was posted to the final day of data collection, September 15, 2020, and multiplied by 1000. n=100 for each cohort.

Variables from the 20 most successful campaigns (based on the calculated CSI) and the bottom 20 campaigns were compared. The top 20 global health campaigns had higher median fundraising goals of $12,500 (IQR: $2,000-$24,590) compared to the bottom 20 (median $5,000). Conversely, the top 20 personal health campaigns had a lower median overall fundraising goal of $20,000 (IQR: $5,625- $33,750) compared to the bottom 20 (median $125,000). This was a similar trend observed in the animal activism campaigns, with a lower median fundraising goal of $7,177 for the top 20 campaigns compared to the bottom 20 campaigns (median $50,000) (Fig 3A). Nonetheless, the top 20 global health campaigns raised a higher median total amount $10,557 (IQR: $2,099-$22,769) compared to the bottom 20 campaigns $2,218 (IQR: $1,230-$4,600). This trend was not observed in the other two cohorts (Fig 3B).

**Fig 3.**
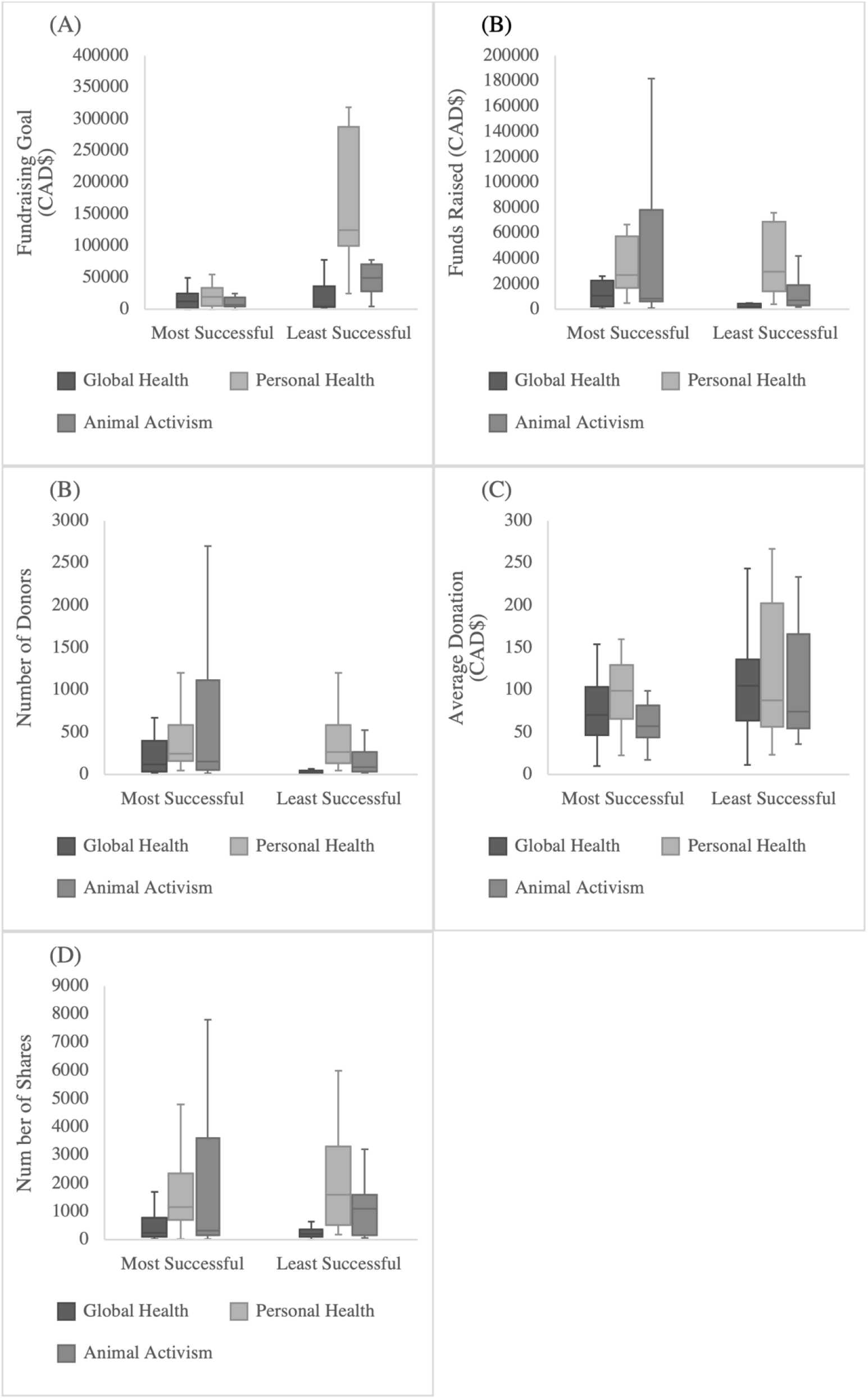
Comparison of factors for campaign success in global health, personal health, and animal activism. Success as defined by campaign success index calculated as the percent of the campaign goal raised divided by number of days from when the campaigned was posted to the final day of data collection, September 15, 2020, and multiplied by 1000.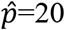 for each cohort. (A) Fundraising goal in CAD$ of the most successful and least successful campaigns. (B) Amount of funds raised in CAD$ by the most successful and least successful campaigns. (C) Number of donors donating to the most successful and least successful campaigns. (D) Average donation amount in CAD$ of the most successful and least successful campaigns, calculated from the funds raised divided by the number of donors to each campaign (n=100) for global health, personal health, and animal activism campaigns. (E) Number of shares of the most successful and least successful campaigns through the GoFundMe share function to social media and content sharing platforms.

Finally, the total number of donors contributing to the top 20 global health and animal activism campaigns were considerably higher than the bottom 20: 119 (IQR: 34-396) compared to 24 (IQR: 14-46) for global health and 151 (IQR: 52-1113) compared to 85 (37-269) for animal activism. In contrast, in personal health crowdfunding drives, the total number of donors were similar between the top 20 and bottom 20 campaigns. However, in these personal health campaigns the average donation amount was higher for the top 20 (Fig 3C). In comparison, the top 20 campaigns for both global and animal had more donors than the bottom 20 but lower average donation amount (Fig 3D). Similarly, the number of social and traditional media shares for global health campaigns were examined, there appeared to be a noteworthy difference with most successful campaigns being more active. The opposite trends were seen in the other cohorts (Fig 3E).

## Discussion

This study utilizing the online crowdfunding site GoFundMe compared the composition, goals, and activities of global health initiatives compared to personal health and animal activism. Several interesting findings were observed including that within the site global health is not a pre-existing campaign category. Of those that were reviewed in this study, nearly half were seeking donation for medical mission trips and a majority organized by the individual going on the mission, consistent with voluntourism. The call for these public donations were mostly for travel expenses or non-specific medical expenses and supplies destined to low resourced populations and only 15% were we able to determine any religious or spiritual affiliation. Compared to personal health initiatives on the platform, which make up approximately one-third of its campaigns (1), global health-based campaigns were dramatically less aspirational in their fundraising goals. Furthermore, the dollars raised were more modest with less engagement compared to personal health and animal activism. Despite the less aggressive global health campaigns with less numbers of donors, they’re relative success was comparable to the other cohorts specifically the average donation amount which appeared equivalent.

Compared to all GoFundMe campaigns from all categories, health-related campaigns have been documented to be more likely to succeed, regardless of the fundraising goal of the campaign, and receive the most donations (9). In this current study, global health campaigns were observed to be the lowest in every measured category except average donation amount: setting the lowest goals, raising the least amount of money, and attracting the least amount of donor attention and shares. Of 100 global health campaigns, the median fundraising goal was $3,000 (IQR: $2,000-$8,500) and raised a median of $2,150 (IQR: $1,600-$4,480) compared to personal health campaigns’ much larger median goal of $52,500 (IQR: $20,000-$150,000) and $32,500 (IQR: $14,250-$75,750) raised. However, in terms of our campaign success index (CSI) relating campaign goals to dollars raised during the length of campaign, global health overall CSI was comparable to those of personal health and animal activism. Successful global health and animal activism campaigns both had more donors, but smaller donation amounts on average than the least successful campaigns, a trend not seen in personal health. This demonstrates that global health and animal activism must attract a larger but less generous audience to be successful compared to personal health.

These results might suggest that these types of online crowdfunding for global health initiatives may be more successful targeting more people donating, understanding that donation amounts will be less. When looking at number of social and traditional media shares for global health campaigns, successful campaigns were clearly more active, interestingly a trend not observed in personal health or animal activism campaigns, suggesting that campaigns raising money for global health endeavours were more reliant on social media or more traditional content sharing platforms. Based on these observations, it is possible that the more modest performance using GoFundMe for global health funding proposals may be a symptom of underutilization rather than an incompatibility of crowdfunding for this purpose. Despite less aspirational funding goals and less engagement with the online public, the average donation amounts received were equivalent to the other campaigns we reviewed. Although global health was the least successful in general, when examining the top performing campaigns, it was apparent that those with higher fundraising goals raised more funds; a trend not found in either personal health or animal activism. This suggests that asking for more generous support lends itself to raising more in regard to global health campaigns, but the same cannot be said for personal health and animal activism campaigns where asking for smaller amounts was more successful.

This study demonstrated a lack of direct funding to international global health initiatives created by those directly needing care. Instead, most campaigns are dedicated to fundraising support for a party to have an experience with the purpose of providing care/service abroad to cover travel expenses for unspecified mission trips thereby demonstrating the profusion of “voluntourism” in crowdfunded global health initiatives. In this sense, it is important to mention that the GoFundMe platform is supported in only 19 countries such that campaigns can only be created within these HICs in North American and Western Europe. This prevents global health initiatives in lower income countries to be self-directed and instead relies on campaigns to be created in higher income countries and extended to other countries in need. Our study was limited by the inability to collect a simple random sample of GoFundMe campaigns due to the website’s uses a complex algorithm to sort the campaigns that are shown to visitors, both in the category sections and via search terms. A GoFundMe charity services support specialist supplied an explanation of the algorithm when contacted: “[the] algorithm calculates the level of engagement with [the] fundraisers.” Such an algorithm may be influenced by social and traditional media attention via the campaign’s share function, as well as the visitor’s IP address to show local “trending” campaigns (10,11). Interestingly, despite the universal health care system enjoyed in Canada none of the global health campaigns were initiated within the country. It is possible that the system in Canada leads to less personal financial need and therefore a differential use of online crowdfunding for health-related endeavors (4).

Limitations of this study include the relatively small sampling of GoFundMe initiatives in each cohort although an iterative process of review by separate study personnel to ensure saturation of some of the qualitative themes facilitated a good reflection of the separate campaigns. However, our data with respect to fundraising goals, raised amount, number of donors, and shares of personal health campaigns were on par to another recent study regarding medical crowdfunding on GoFundMe (4). GoFundMe provides no explicit global health category in their search function so campaigns were identified from general search terms which might have led to some sampling bias. As well, there are inherent limitations in manual review and veracity of the information supplied within the individual webpages. Generalizability of our findings is limited by the use of a single crowdsourcing platform. Watsi (https://watsi.org/) is another crowdfunding website that takes a different approach to charitable crowdfunding for individuals in LMICs countries, connecting donors to patients already seeking treatment at partnered medical entities or NGOs. Watsi actively selects which campaigns are approved, has the posts created by writing staff, and handles all transfers of funds to the medical entity in an attempt to promote only authentic cases. Watsi is also known for its use of radical transparency to uphold authenticity by posting an annual spreadsheet of all campaign data including patients, medical partners, dollar amounts, and outcomes. However, some have questioned such methods for prioritizing high impact or high success treatments over palliative or low success treatments and offering them to a public audience (12). Further work is needed to explore the trends in this study, extending these observations on other crowdfunding platforms, in order to determine factors affecting funding success as well as understand the social and ethical implications of online solicitation for global health endeavors.

## Data Availability

All data is included in the submission

## S1 Text. Counties Supported by GoFundMe Platform

GoFundMe lists 19 countries on their website in which the platform is supported as of 2020: Australia, Austria, Belgium, Canada, Denmark, Finland, France, Germany, Ireland, Italy, Luxembourg, Netherlands, Norway, Portugal, Spain, Sweden, Switzerland, United Kingdom, and the United States excluding American Samoa, Guam, Northern Mariana Islands, Puerto Rico, and U.S. Virgin Islands (https://support.gofundme.com/hc/en-us/articles/360001972748-What-Countries-are-Supported-on-GoFundMe-).

